# Anti-SARS-CoV-2 Serology persistence over time in COVID-19 Convalescent Plasma Donors

**DOI:** 10.1101/2021.03.08.21253093

**Authors:** Valeria De Giorgi, Kamille A West, Amanda N Henning, Leonard Chen, Michael R Holbrook, Robin Gross, Janie Liang, Elena Postnikova, Joni Trenbeath, Sarah Pogue, Tania Scinto, Harvey J Alter, Cathy Corny Cantilena

## Abstract

**Background:** Characterizing the kinetics of the antibody response to SARS□CoV□2 is of critical importance to developing strategies that may mitigate the public health burden of COVID-19. We sought to determine how circulating antibody levels change over time following natural infection.

**Methods/Materials:** We conducted a prospective, longitudinal analysis of COVID-19 convalescent plasma (CCP) donors at multiple time points over a 9-month period. At each study visit, subjects either donated plasma or only had study samples drawn. In all cases, anti-SARS-CoV-2 donor testing was performed using semi-quantitative chemiluminescent immunoassays (ChLIA) targeting subunit 1 (S1) of the SARS-CoV-2 spike (S) protein, and an in-house fluorescence reduction neutralization assay (FRNA).

**Results:** From April to November 2020 we enrolled 202 donors, mean age 47.3 ±14.7 years, 55% female, 75% Caucasian. Most donors reported a mild clinical course (91%, n=171) without hospitalization. One hundred and five (105) (52%) donors presented for repeat visits with a median 42 (12-163) days between visits. The final visit occurred at a median 160 (53-273) days post-symptom resolution. Total anti-SARS-CoV-2 antibodies (Ab), SARS-CoV-2 specific IgG and neutralizing antibodies were detected in 97.5%, 91.1%, and 74% of donors respectively at initial presentation. Neutralizing Ab titers based on FRNA_50_ were positively associated with mean IgG levels (p = <0.0001). Mean IgG levels and neutralizing titers were positively associated with COVID-19 severity, increased donor age and BMI (p=0.0006 and p=0.0028, p=0.0083 and p=0.0363, (p=0.0008 and p=0.0018, respectively). Over the course of repeat visits, IgG decreased in 74.1% of donors; FRNA_50_ decreased in 44.4% and remained unchanged in 33.3% of repeat donors. A weak negative correlation was observed between total Ab levels and number of days post-symptom recovery (r = 0.09).

**Conclusion:** Anti-SARS-CoV-2 antibodies were identified in 97% of convalescent donors at initial presentation. In a cohort that largely did not require hospitalization. IgG and neutralizing antibodies were positively correlated with age, BMI and clinical severity, and persisted for up to 9 months post-recovery from natural infection. On repeat presentation, IgG anti-SARS-CoV-2 levels decreased in 56% of repeat donors. Overall, these data suggest that CP donors possess a wide range of IgG and neutralizing antibody levels that are proportionally distributed across demographics, with the exception of age, BMI and clinical severity.

## Introduction

The ongoing COVID-19 pandemic, caused by the novel coronavirus severe acute respiratory syndrome coronavirus 2 (SARS-CoV-2), constitutes a global health crisis with devastating social and economic consequences. As efforts are underway to curtail further spread of COVID-19 worldwide, it is critical to understand the longevity and potency of immune response to SARS□CoV□2. Antibody production represents a significant component of the immune response to SARS-CoV-2, and serologic assays to detect circulating antibody levels are a readily available tool in clinical laboratory settings for tracking immune responses over time [1, 2].

Due to the recent emergence of this infectious disease, there is a relative paucity of data on the long-term kinetics of SARS-CoV-2 antibodies. Studies of individuals who have recovered from natural infection may help to determine how long antibodies persist after an immunizing exposure, and whether or not these antibodies might protect against re-infection. The persistence of antibody response may also help predict the efficacy of vaccines for COVID□19.

COVID-19 convalescent plasma (CCP) is an investigational therapy that remains somewhat controversial, with conflicting reports of efficacy [3, 4, 5]. The underlying principle of this treatment approach is the passive transfer of anti-SARS-CoV-2 antibodies to patients with COVID-19. Serologic assays therefore play a critical role in identifying suitable convalescent plasma donors, and provide evidence of humoral immunity against SARS-CoV-2.

Importantly, as CCP donors may return for multiple repeat donations, these individuals provide a valuable opportunity to characterize anti-SARS-CoV-2 serologic responses and to determine how circulating antibody levels change over time in a well-defined cohort [6].

Of the four major structural proteins for SARS-Co-V2, spike (S), membrane (M), envelope (E), and nucleocapsid (N) proteins, the S and the N proteins are both highly immunogenic and abundantly expressed during the infection [7]. They have been used as target antigens for the majority of serological assays. The S glycoprotein, however, is surface-exposed and mediates entry into host cells. As such it is subject to a more stringent selection pressure from the immune system, and for this reason, it represents the main target of neutralizing antibodies [8].

In the current study, we prospectively conducted a longitudinal serological assessment of CCP donors after recovery from natural infection. Donors were assessed for levels of neutralizing antibodies as well as total and IgG-specific S protein antibodies using a lab developed plaque reduction assay (FRNA) and a commercially available system (Ortho VITROS®). Data were analyzed to identify correlations between antibody levels and clinical characteristics, and we present follow-up serological data in repeat CCP donors. Collectively, we present a comprehensive view of SARS-CoV-2 antibody dynamics over a 9-month period after natural infection, increasing our understanding of COVID-19 immune responses among persons with community acquired SARS-CoV-2 infection.

## Methods

### CCP Donor qualification and plasma collection

COVID-19 convalescent plasma donors were prospectively enrolled onto an IRB-approved protocol (Clinical Trials Number: NCT04360278) and provided written informed consent for the study. Eligibility criteria included (1) routine blood donor criteria, (2) molecular or serologic laboratory evidence of past COVID-19 infection and (3) complete recovery from COVID-19, with no symptoms for ≥ 28 days, or ≥ 14 days with a negative molecular test after recovery.

We collected donor demographic and biometric data, including age, race, sex and body mass index. We categorized clinical severity of past COVID-19 infection as asymptomatic, mild (self-limiting course, symptomatic management at home), moderate (emergency room management or hospitalization) or severe (ICU admission). Depending on center-specific logistics and changes in demand for convalescent plasma, at each study visit, subjects either donated plasma, or only had whole blood samples drawn for research; in all cases, anti-SARS-CoV-2 testing was performed. Based on the results of the previous visit, donors were recruited to either continue donating plasma, or donate samples for research purposes. Plasma collections were performed by apheresis, yielding 600 – 825 mL of donor plasma. Sample draws were limited to < 70 mL of whole blood per visit. The minimum interval between plasma donations was 28 days; shorter intervals were acceptable between sample draw visits. Routine plasma donor testing was performed, including standard infectious disease testing, blood group assessment and HLA antibody testing in female donors.

Plasma for anti-SARS-CoV-2 testing was isolated from EDTA-anticoagulated human whole blood samples. Samples were processed immediately after collection, centrifuged for 15 min at 2500rpm to separate the plasma phase. The upper plasma layer was extracted, aliquoted and stored at −80C in our research repository after antibody testing and then shipped on dry ice to the NIH/National Institute of Allergy and Infectious Diseases (NIAID) Integrated Research Facility at Fort Detrick, Maryland, USA, for the in-house assessment of the sample’s SARS-CoV-2 neutralization activity.

Prior to August 2020, the FDA recommended titers of at least 1:160 for investigational convalescent plasma, but a titer of 1:80 was considered acceptable if higher-titer units were not available. In August 2020, the FDA granted COVID-19 convalescent plasma emergency use authorization; the criteria for high-titer CCP have subsequently been revised several times.

### SARS-CoV-2 Immunoassay

Donor samples were tested for SARS-CoV-2 antibodies using the Ortho-Clinical Diagnostics VITROS^®^ Total (IgA/G/M) and IgG COVID-19 Antibody tests following the manufacturer’s package insert instructions (https://www.fda.gov/media/136967/download). Both assays have been granted Emergency Use Authorization (EUA) by the FDA. The Ortho-Clinical Diagnostics anti-SARS-CoV-2 assay is a qualitative CLIA utilizing luminol-horseradish peroxidase (HRP)- mediated chemiluminescence assay, performed on the VITROS^®^ 3600 automated immunoassay analyzer. The assay involves a two-stage reaction. In the first stage antibodies to SARS-CoV-2 present in the sample bind with SARS-CoV-2, spike protein coated on wells. Any unbound sample is removed by washing. In the second stage, HRP-labeled murine monoclonal anti-human IgG antibodies (or recombinant SARS-CoV-2 spike protein S1 antigen for the total assay) are added in the conjugate reagent. The conjugate binds specifically to the antibody portion of the antigen-antibody complex. If complexes are not present, the unbound conjugate is removed by the subsequent washing step. The HRP in the bound conjugate catalyzed oxidation of the luminol, which produced a chemiluminescent reaction measured by a luminometer. The patient sample signal is divided by the calibrator signal, with the resulting S/Co (Signal/Cutoff) values of <1.00 and ≥1.00 corresponding to nonreactive and reactive results, respectively.

### SARS-CoV-2 Neutralizing assay

Assays for determining neutralizing titers were performed with authentic SARS-CoV-2 (2019-nCoV/USA-WA1-A12/2020 from the US Centers for Disease Control and Prevention, Atlanta, GA, USA) at the NIH-NIAID Integrated Research Facility at Fort Detrick, MD, USA using a fluorescence reduction neutralization assay (FRNA). This assay was performed by incubating a fixed volume of virus (0.5 multiplicity of infection (MOI)) with an equivalent volume of test plasma for 1 h at 37 ºC prior to adding to Vero E6 cells (BEI, Manassas, VA, USA, #NR-596) plated in 96-well plates. Following addition to Vero E6 cells, the virus was allowed to infect the cells and propagate for 24 hours at 37 ºC/5% CO_2_, at which time the cells were fixed with neutral buffered formalin. Following fixation, the cells were permeabilized with RIPA buffer and probed with a SARS-CoV-2 nucleoprotein-specific rabbit primary antibody (Sino Biological, Wayne, PA, USA, #40143-R001) followed by an Alexa647-conjugated secondary antibody (Life Technologies, San Diego, CA, USA, #A21245). Cells were counterstained with Hoechst nuclear stain (Life Technologies #H3570). Cells in four fields per well were counted with each field containing at least 1000 cells, with four wells per dilution step for each test sample. Plates were read and quantified using an Operetta high content imaging system (PerkinElmer, Waltham, MA). Antibodies were screened using a 2-fold serial 6-step dilution. Results are reported as the highest dilution of plasma leading to at least 50% reduction in SARS-CoV-2 titers (FRNA_50_) when compared to virus only controls. The lower limit of detection was 1:40. Assays were controlled using a spike protein specific antibody as positive control in addition to virus-only and uninfected cell controls [9]. All data were analyzed using Excel. To detect potential experimental errors, we used Dixon’s Q-test, a quick statistical method to detect gross errors in small samples. We used the critical value of 0.829 for 95% confidence level at N=4 [10, 11].

### Statistical Analysis

All statistical analyses were performed using GraphPad Prism version 8.4.3 (www.graphpad.com). Relationship between multiple groups were examined using the non-parametric Kruskal-Wallis test, with post hoc comparisons calculated using Dunn’s multiple comparison test. US population demographic data for age and sex were obtained from the 2019 US Census Bureau survey (www.census.gov). Blood type frequencies were obtained from Garratty et al. [12].

## Results

### COVID-19 Convalescent Plasma Donor Characteristics

We enrolled 202 COVID-19 convalescent plasma (CCP) donors between April and November of 2020. The median age of the cohort was 47 (19-79) years, 54.5% of donors were female. The distribution of donor ABO/Rh blood groups was as follow: A+ (29.7%), A-(5.9%), AB+ (6.9%), AB-(0.5%), B+ (12.4%), B-(0.5%), O+ (33.7%) and O-(5.4%), which was not statistically different from the US general population ^(11,12)^ (Fig 1a-c). HLA antibodies were found in 24% of females. The overwhelming majority of donors reported a mild disease course (182, 90%). Most donors self-identified as Caucasian (75.7%), Asian (7.4%), Black (6.9%), or Hispanic (4.5%). Additional CCP donor characteristics are shown in Table 1.

**Table 1.** CCP Donor Characteristics.

|  | All Donors<br>(n=202) | Repeat Donors<br>(n=105) |
| --- | --- | --- |
| Sex | N (%) | N (%) |
| Female | 110 (54.5) | 58 (55.2) |
| Male | 92 (45.5) | 47 (44.8) |
| Disease Course* | N (%) | N (%) |
| Asymptomatic | 3 (1.5) | 2 (1.9) |
| Mild | 182 (90.1) | 94 (89.5) |
| Moderate | 14 (6.9) | 7 (6.7) |
| Severe | 3 (1.5) | 2 (1.9) |
| Race | N (%) | N (%) |
| Asian | 15 (7.4) | 8 (7.6) |
| Black | 14 (6.9) | 4 (3.8) |
| Caucasian | 153 (75.7) | 79 (75.2) |
| Hispanic | 9 (4.5) | 7 (6.7) |
| Mixed Race | 6 (3.0) | 4 (3.8) |
| Other/Unknown | 3 (1.5) | 2 (1.9) |
| Declined | 2 (1.0) | 1 (1.0) |
| Blood Type | N (%) | N (%) |
| A- | 12 (5.9) | 6 (5.7) |
| A+ | 60 (29.7) | 31 (29.5) |
| AB- | 1 (0.5) | 1 (1.0) |
| AB+ | 14 (6.9) | 8 (7.6) |
| B- | 1 (0.5) | 1 (1.0) |
| B+ | 25 (12.4) | 15 (14.3) |
| O- | 11 (5.4) | 5 (4.8) |
| O+ | 68 (33.7) | 38 (36.2) |
| Unknown | 10 (5.0) | - |
| HLA status in Females | N (%) | N (%) |
| Negative | 81 (73.6) | 44 (75.9) |
| Positive | 26 (23.6) | 13 (22.4) |
| Unknown | 3 (2.7) | 1 (1.7) |
|  | Median (Range) | Median (Range) |
| Age | 47 (19-79) | 50 (19-79) |
| BMI | 26.0 (17.9-51.8) | 25.9 (17.9-51.8) |

| Days post-symptom resolution | Median (Range) | Median (Range) |
| --- | --- | --- |
| 1st donation | 50 (18-244) | 43 (18-244) |
| 2nd donation | - | 106 (53-273) |
| 3rd donation | - | 145 (88-248) |
| 4th donation | - | 159.5 (122-245) |
| 5th donation | - | 205.5 (157-224) |
| 6th donation | - | 196 (187-204) |
| 7th donation | - | 224 (216-232) |
| Final donation | - | 160 (53-273) |

| Days between | Median (Range) | Median (Range) |
| --- | --- | --- |
| Consecutive donations | - | 42 (12-163) |
| 1st and final donation | - | 90 (21-206) |

| # of Donations | N (%) | N (%) |
| --- | --- | --- |
| 2 donation | - | 47 (44.8) |
| 3 donations | - | 36 (34.3) |
| 4 donations | - | 8 (7.6) |
| 5 donations | - | 11 (10.5) |
| 6 donations | - | 1 (1.0) |
| 7 donations | - | 2 (1.9) |

\***mild**= self-limiting course, symptomatic management at home; **moderate**= emergency room management or hospitalization; **severe**= ICU admission

**Fig. 1.**
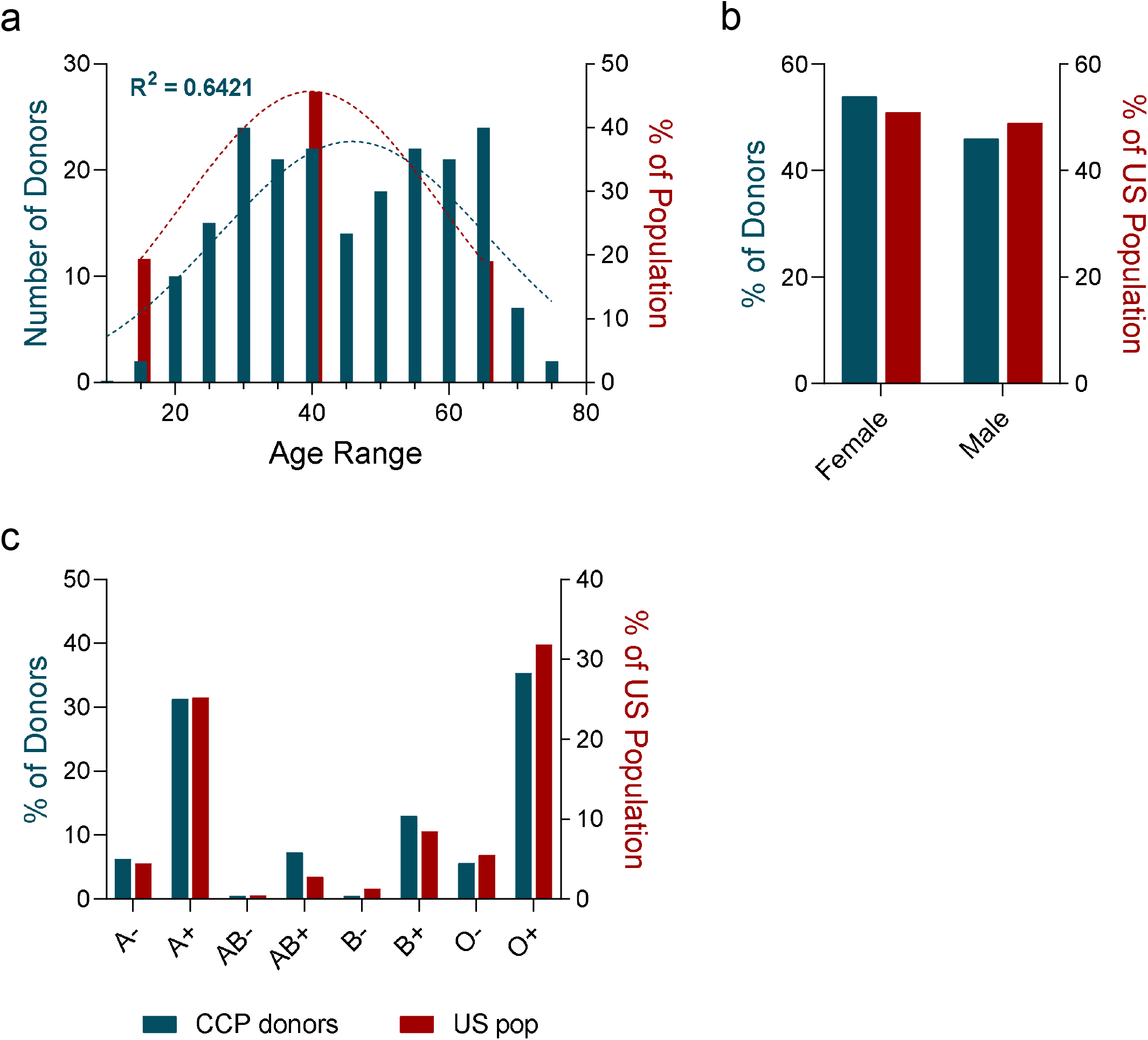
COVID-19 convalescent plasma donor characteristics. **a.** Age distribution of COVID-19 convalescent plasma (CCP) donors (blue; n=202) relative to the US population (red). Dotted lines represent a Gaussian distribution and the coefficient of determination (R^2^) was calculated for the CCP goodness of fit. **b**. Sex distribution of CCP donors (blue; n=202) relative to the US population (red). Binomial test not significant relative to the US population. **c**. Blood type distribution of CCP donors (blue; n=192) relative to the US population (red). Binomial tests performed relative to US population, all comparisons not significant following Bonferroni correction for multiple comparisons.

### SARS-CoV-2 Seroconversion and Neutralizing Activity at Initial Presentation

At first study visit, donors presented at a median of 50 (18-244) days post-symptom resolution. Total SARS-CoV-2 antibodies were detected in 97.5% of donors (197/202) and IgG antibodies detected in 91.1% of donors (184/202) (Fig.2a). The median S/Co values of CCP plasma samples using the Ortho total Ab and Ortho IgG assay were 203.5 (0.1-969) and 8.5 (0.01-30.7), respectively. In 46.5 % (94/202) of the donors IgG antibody levels at initial presentation met the current FDA EUA high titer criterion of S/Co ≥ 9.5 when tested with IgG VITROS®. Total SARS-CoV-2 antibody levels were strongly correlated with IgG antibody levels (r = 0.7814, p = <0.0001) (Fig 2b). Neutralizing activity as measured by FRNA_50_ was observed in 73.8% of samples (149/202) (Fig 2a). The majority of donors (173, 85.6%) had relatively modest neutralizing activity (FRNA_50_ titer ≤1:80). Specifically 124 (61.4%) had an FRNA_50_ titer of <1:80, and 49 (24.3%) donors had a titer of 1:80, which prior to August 2020 was the minimum acceptable neutralizing antibody titer for CCP per FDA recommendations. A small proportion of CCP donors had higher neutralizing activity (FRNA_50_ titer >1:80), 17 (8.4%) had an FRNA_50_ titer of 1:160, 9 (4.5%) had a titer of 1:320, and 3 (1.5%) had a titer of 1:640. In our cohort, a statistically significant difference in mean IgG levels among neutralizing titer groups was observed (Kruskal-Wallis test; *H* (6) = 97.84, p = <0.0001). Specifically, average IgG antibody levels were significantly greater in samples with high FRNA_50_ titer (1:80 or higher) than in low titer samples (<1:80) (Fig 2c), suggesting a positive correlation between IgG antibodies and SARS-CoV-2 neutralizing activity. We observed weak correlations between total antibody levels or IgG antibody levels and duration of convalescence (expressed as days post-symptom resolution). Specifically, over time total antibodies displayed a weak but significant positive correlation (r = 0.2676, p = 0.0001), while IgG antibodies displayed a weak negative correlation of borderline significance (r = −0.1403, p = 0.0481) (Extended data Fig 1a). A statistically significant difference in mean days post-symptom resolution and neutralizing titer was observed (Kruskal-Wallis test; *H* (6) = 19.60, p = 0.0033), with post hoc pairwise comparisons suggesting a weak negative correlation between the two (Extended Data Fig. 1b).

**Fig. 2.**
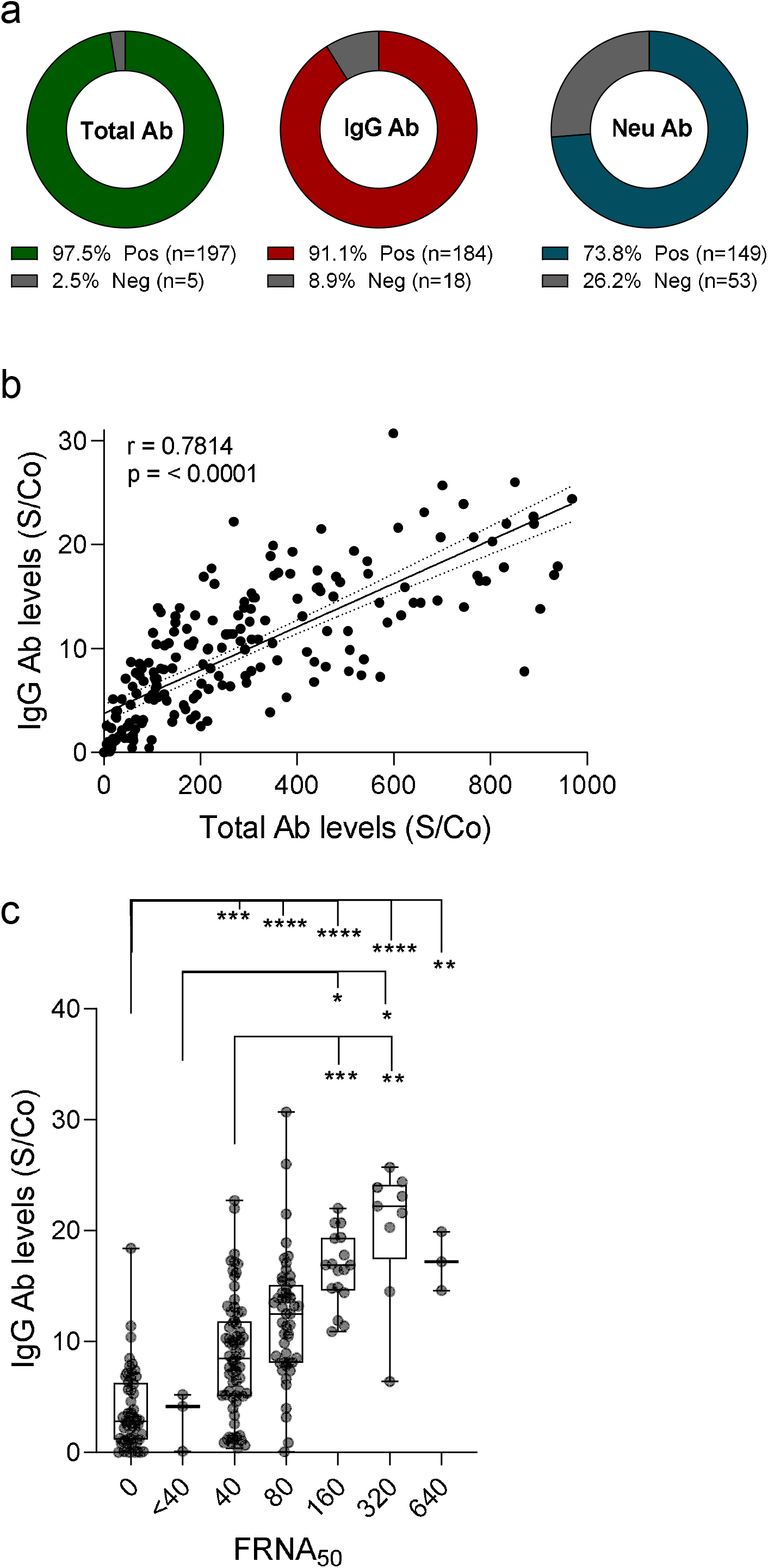
SARS-CoV-2 seroconversion and neutralizing activity at initial donation. **a.** Percent of CCP donors testing positive (Pos) or negative (Neg) for the presence of total antibodies (Total Ab), IgG antibodies (IgG Ab), or neutralizing antibodies (Neu Ab). **b**. Correlation of total antibody levels with IgG antibody levels (n=202). Line represents a linear fit of the data including 95% CI (dotted lines). Pearson correlation coefficient (r) is displayed along with p-value. **c**. Distribution of IgG antibody levels based on FRNA_50_ neutralizing antibody titers (n=202). Boxplots indicate median value with 1^st^ and 3^rd^ quartiles, and bars span minimum and maximum values. Individual patient values are indicated by dots. A statistically significant difference between groups was determined via Kruskal-Wallis test (p-value <0.0001). Post hoc comparisons were calculated using Dunn’s multiple comparison test, with significant pairwise comparisons indicated on the graph. S/Co, signal to cutoff ratio; *, p <0.05; **, p <0.01; ***, p <0.001; ****, p <0.0001.

### Correlation of CCP donor Serology Tests and Neutralizing Activity with Clinical Characteristics

A severe clinical COVID-19 course requiring ICU admission, was associated with increased levels of total and IgG antibodies at initial presentation (p=0.0006 and p=0.0028, respectively). There was a trend toward association between COVID disease severity and neutralizing antibody titer, which did not achieve statistical significance (p=0.0527) (Fig 3a). Increased IgG antibody levels and neutralizing titers were associated with both increased donor age (p=0.0083 and p=0.0363, respectively) and BMI (p=0.0008 and p=0.0018, respectively) (Fig 3b-c). We observed no associations between total antibody levels, IgG levels, or neutralizing antibody titers and sex, HLA status in females, blood type, or race (Extended Data Fig1c-f).

**Fig. 3.**
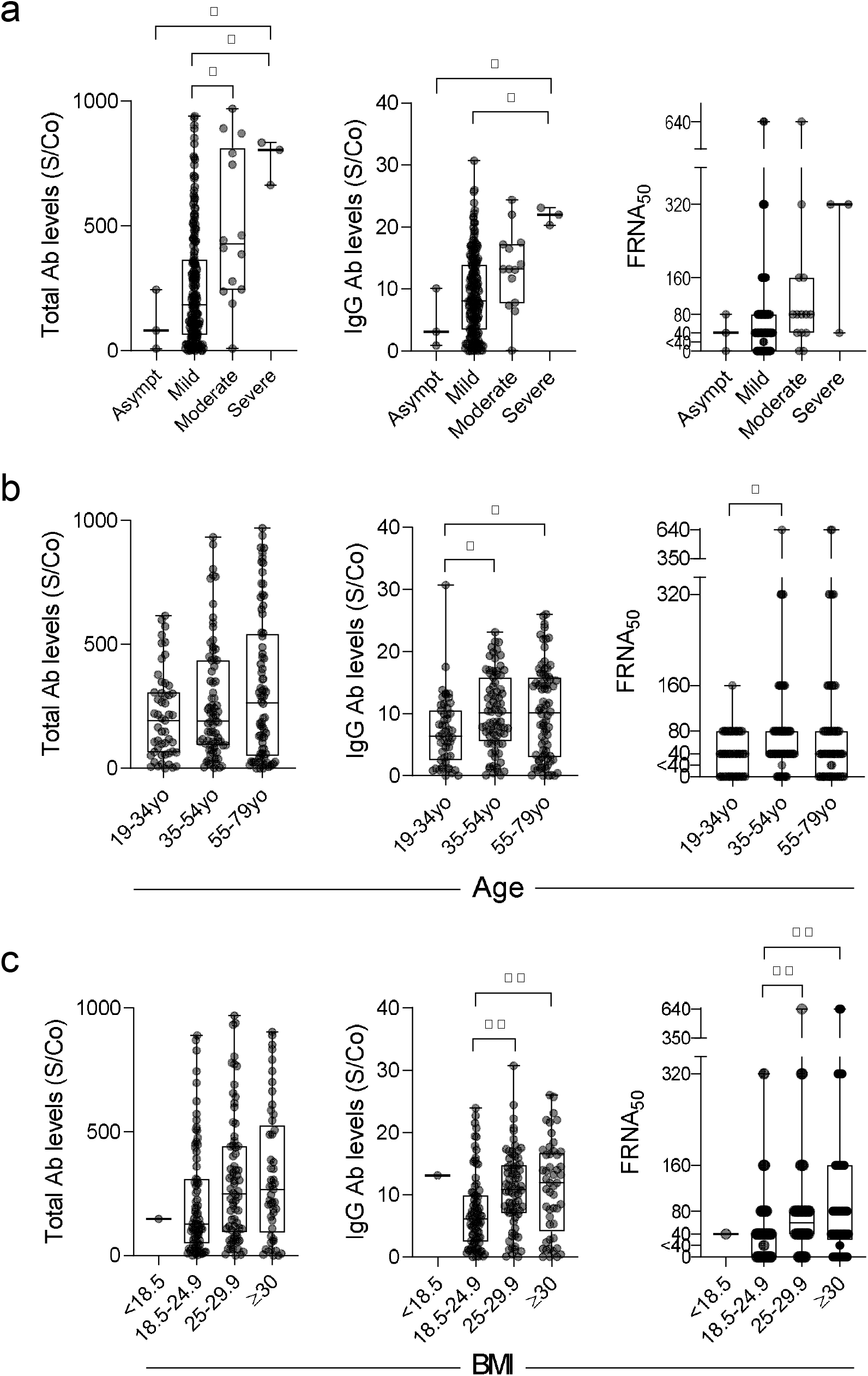
Correlation of serology tests and neutralizing activity with clinical characteristics. Total antibody levels, IgG antibody levels, and neutralizing antibody titers (FRNA_50_) in CCP donors are shown based on **a**. clinical disease course (asymptomatic, n=3; mild, n=182; moderate, n=14; severe, n=3), **b**. donor age (19-34yo, n=51; 35-54yo, n=75; 55-79yo, n=76), and **c**. donor BMI (<18.5, n=1; 18.5-24.9, n=79; 25-29.9, n=72; ≥30, n=49; missing data for n=1 donor). For all graphs, boxplots display the median with 1^st^ and 3^rd^ quartiles, and bars span minimum and maximum values. Individual patient values are indicated by dots. Statistically significant differences between groups were determined via Kruskal-Wallis test for total ab levels and IgG ab levels based on disease course (p = 0.0006 and 0.0028, respectively) (**a**), for IgG ab levels and neutralizing titers based on age (p-value = 0.0083 and 0.0363, respectively) (**b**), and for IgG ab levels and neutralizing titers based on BMI (p-value = 0.0008 and 0.0018, respectively) (**c**). Post hoc comparisons were calculated using Dunn’s multiple comparison test, with significant pairwise comparisons indicated on graphs. *, p <0.05; **, p <0.01.

### SARS-CoV-2 Antibody Levels in CCP Donors over Time

105/202 (52%) plasma donors returned for repeat study visits, allowing for IgG and neutralizing antibody analyses at multiple time points. Repeat donor demographics were similar to our overall cohort. Most repeat donors presented 2 (44.8%) or 3 (4.3%) times, 20.9% (n = 22/105) presented for 4 or more study visits (Table1). The median interval between consecutive visits was 42 (12-163) days and between the first and final visits was 90 (21-206) days (Fig 4a). Across all repeat donors, the first visit time point occurred at a median of 43 (18-244) days post-symptom resolution, and the final time point occurred at a median of 160 (53-273) days post-symptom resolution (Fig 4b).

**Fig. 4.**
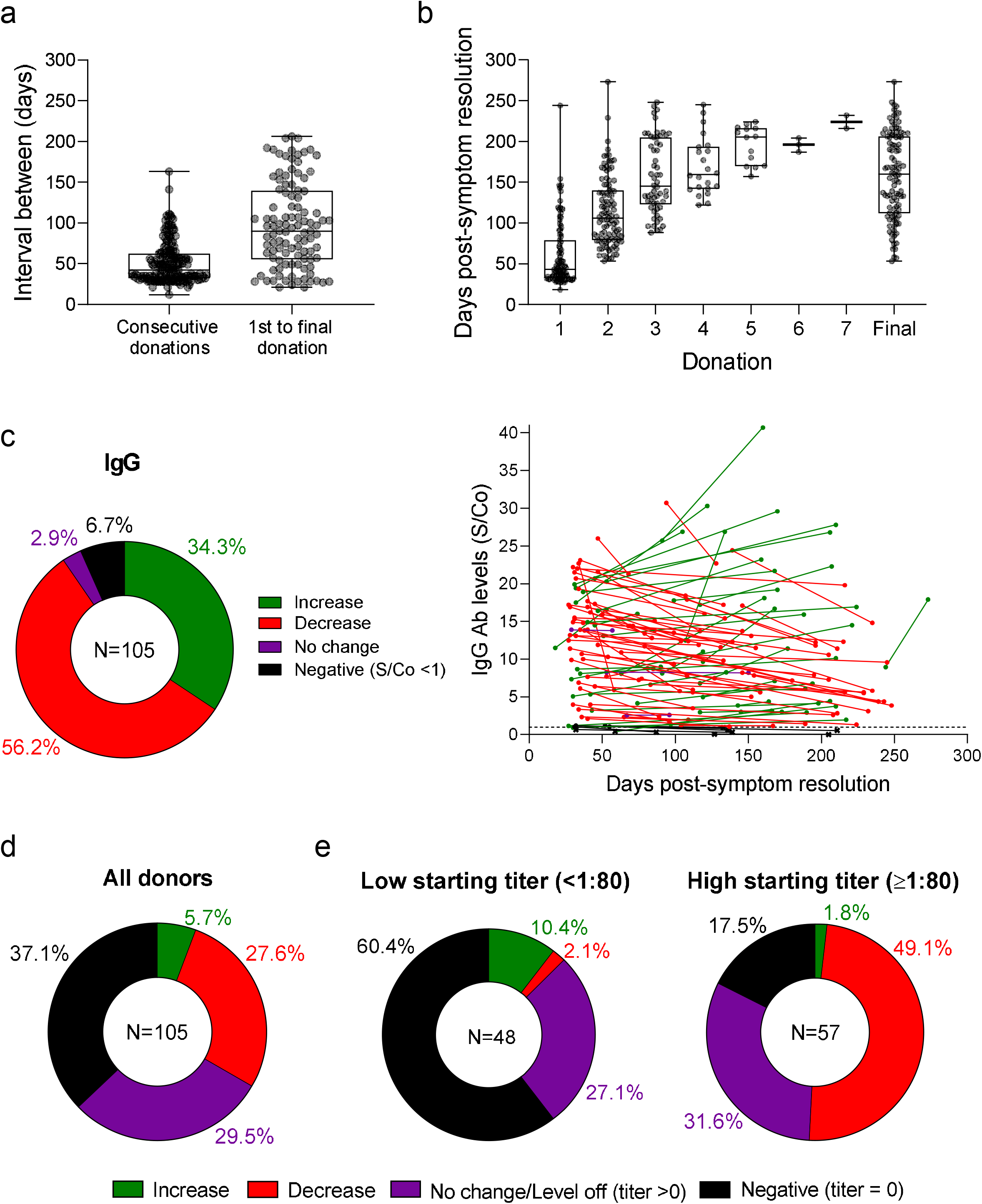
SARS-CoV-2 antibody levels in CCP donors over time. **a.** Boxplots indicate the interval between consecutive donations and between first and final donation. **b**. Boxplots display median number of days post-symptom resolution at indicated donations. For **a** and **b**, boxplots indicate median value with 1^st^ and 3^rd^ quartiles, and bars span minimum and maximum values. Individual patient values indicated by dots. **c**. IgG antibody outcomes over time for repeat CCP donors are quantified in the pie chart, and individual antibody levels are plotted relative to days post-symptom resolution (n=103; n=2 asymptomatic donors not plotted). Only first and last CCP donation values are plotted. Samples from the same donor connected by lines. Colors indicate antibody level outcomes over time: increase (green), decrease (red), no change (percent change <1%; purple), and negative (levels consistently below or falling below cutoff threshold S/Co <1; black). The S/Co threshold of 1 indicated by the dashed line. Values below this threshold are represented by an “x”. **d**. Neutralizing titer outcomes over time for repeat CCP donors are quantified. Outcome categories are the same as in **c**, except: No change/Level off indicates donors whose titers remained unchanged or leveled off to a titer >0 for their last 2 or more donations, and Negative indicates donors with consistently undetectable activity or whose titers fell to an undetectable level. **e**. Outcomes from **d** stratified based on the titer at first donation. S/Co, signal to cutoff ratio.

IgG levels decreased over time in a majority of repeat donors (56.2%, 59/105) but increased in 34.3% (36/105) of donors. IgG levels remained unchanged in 2.9% of donors (3/105). In 6.7% of CCP donors (7/105), IgG levels fell below or remained consistently below the cutoff threshold (S/Co < 1) (Fig 4c, Extended Data Fig 2a). Importantly, of the 7 donors with undetectable IgG levels at their final visit, 3 had undetectable IgG levels at initial presentation and the remaining 4 had very low initial IgG S/Co values (S/Co < 2). For donors whose IgG levels decreased over time, an average decrease of 30.7% ± 18.8% in the S/Co value was observed between the first and final visit. For donors whose IgG levels increased over time, the mean increase in IgG S/Co was 56.6% ± 49.8%.

In our analysis of SARS-CoV-2 neutralizing antibody titers over time, for 27.6% (29/105) of repeat CCP donors the FRNA_50_ titers decreased, and increased in only 5.7% (6/105) of donors.

In 29.5% (31/105) of donors, neutralizing antibody titers remained unchanged over time or leveled off to a detectable titer for two or more consecutive final time points. Importantly, 37.1% (39/105) of donors, had no detectable neutralizing activity over time (n=13) or had titers fall to undetectable levels (n=26) (Fig 4d). We observed a striking relationship between neutralizing titer levels at first assessment and titer outcome over time. In general, donors with lower initial FRNA_50_ titers (<1:80) were more likely to lose neutralizing activity over time, while donors with higher starting titers were more likely to have their titer levels remain constant or level off over time (Extended Data Fig 2b). This can be seen when repeat CCP donors are stratified based on low (< 1:80) or high (> 1:80) starting titer levels at the study visit. For CCP donors with low initial neutralizing titer, 60.4% of donors (29/48) lost neutralizing activity or had titers that remained undetectable over time, compared to 17.5% (10/57) of high titer CCP donors. Conversely, 31.6% of high neutralizing titer donors (18/57) compared to 27.1% of low titer donors (13/48) had titers that remained unchanged or leveled off over time (Fig 4e). Additionally, in the high neutralizing titer group, the majority of donors saw their titer levels decrease over time (49.1%, 28/57). It remains to be seen whether titers would continue to fall for these donors, or would level off over additional time points. Of note, no SARS-CoV-2 re-infection was observed in the study cohort.

## Discussion

SARS-CoV-2 has infected millions of individuals globally and, as of February 2021, has led to the death of > 2 million individuals (https://coronavirus.jhu.edu) worldwide. One of the reason for the explosive increase in cases is the limited pre-existing immunity to the virus. As the vaccination, effort around the world began with multiple vaccine types and candidates, evolving variants add complexity to the plan to outrun the virus by vaccination. Cases of re-infections began to be reported in late 2020 [13, 14, 15]. Most of the reported re-infections were far less severe than the first infection in those early reports, boosting the optimistic hypothesis that immune memory of the millions COVID-19 survivors would be able to contribute to the herd immunity to tame the pandemic. However, Zucman et al. described a somber case that is an exception to those mild reinfections described to date, reporting a more severe re-infection caused by the South African variant 20H/501Y.V2 (B.1.351) [16].

As shown from studies of common human coronaviruses, neutralizing antibodies are induced and can last for years, providing protection from re-infection or, in some cases, from aggressive disease [17]. Less definitive data are available for SARS-CoV-2, and the emergence of multiple viral variants and the potential for more severe re-infection underscores the need to better understand the body’s natural immune response to SARS-CoV-2. Undoubtedly, a better understanding of antibody kinetics and protective immunity will be critical in terms of protection against reinfection, for public health policy and vaccine development for COVID-19. To begin to address these questions, we conducted a prospective, longitudinal analysis of 202 COVID-19 convalescent plasma (CCP) donors status with multiple time points over a 9-month period. Studying recovered individuals provides an important opportunity to investigate both antiviral antibody quantification and population immunity conferred after exposure to SARS-CoV-2. Additionally, having repeat serological assessments allowed for the investigation of how circulating antibody levels change over time following natural infection.

At initial assessment, total and IgG SARS-CoV-2 antibodies were detected in 97.5% and 74.3% of donors respectively. Total SARS-CoV-2 antibody levels were strongly correlated with IgG antibody levels, and neutralizing activity was observed in 73.8% of donors.

Our study suggests that most CCP plasma donors, regardless of COVID19 disease severity, have antibodies to SARS-CoV-2 and more than 70% possess neutralizing antibodies. Notably and in accordance with already published studies, we have observed a considerable range in antibody titers in recovered COVID-19 donors [18-19].

When correlating IgG antibody levels and neutralizing titers with clinical picture, our results are consistent with previously published data, observing positive correlation with disease severity [20-23], age [24-25], and BMI [26-27]. While some reports observed a correlation between antibody levels and male sex [28], we did not observe a statistically significant association in our study. This may be due to the predominantly mild symptomatic nature of our cohort, as males are also more likely to develop severe COVID-19 [29], presenting a potential confounding factor.

Longitudinal analyses of 105 CCP donors revealed that 93.3% of donors had detectable IgG levels 9 months after recovery. Although the majority of repeat donors demonstrated a decrease in IgG levels over follow up, IgG levels decreased by, only 30.7% on average, and seroreversion to negative was uncommon. Indeed, of the 6.7% of donors with undetectable IgG levels at their final study visit, just over half of these had no detectible IgG at the initial collection, while the rest represented cases of seroreversion (overall seroreversion rate of 3.8%). It is unlikely that the decline in IgG level was related to the number or frequency of plasma donations in this cohort, since in general, IgG levels do not appreciably decrease over the course of a few months of infrequent plasma donation [30].

With respect to neutralizing antibody, SARS-CoV-2 neutralizing antibody levels tended to remain detectable, irrespective of the disease severity. We found that 63% of donors had detectable neutralizing titers up to 9 months post-recovery, with 25% of donors seroreverting, and 12% of donors failing to make detectible neutralizing antibody titers over the course of the study. Although we have observed a decrease in plasma neutralizing activity over time, neutralizing antibody titers remained measurable in a majority of donors.

Importantly, we observed that if donors were stratified based on their titer level at initial assessment, 82.5% of donors with high starting titer (>1:80) maintained detectible titers over time, while this dropped to 39.6% for those with low starting titer (<1:80). Therefore, our data suggest that neutralizing activity in the early convalescent period may be predictive of long-term neutralizing activity retention.

Our results support sustained immunological memory for most of the first year following SARS-CoV-2 infection, even for mild COVID-19 cases, consistent with other reports [31-33]. Furthermore, our data suggest that circulating IgG and neutralizing antibodies persist in most healthy people over the first year following symptom resolution, and longer-term follow-up studies by others have observed similar results out to 8 months [34-38]. However, other reports have observed a decline in immunological response over the first months following symptom resolution, particularly in mild and asymptomatic cases [39-41]. Such discordances could have major implications for modelling of disease transmission, herd immunity and vaccine strategies. Moving forward, specific consideration should be given to the selection of the SARS-CoV-2 antibody assays in order to have seroprevalence estimates that are comparable and devise standardized approaches (e.g. antigen target selection, time of testing, assay sensitivity and specificity, role of IgA).

The levels of neutralizing antibody activity required to prevent SARS-CoV-2 re-infection are currently unknown. In our cohort, no SARS-CoV-2 re-infection was identified. However, we observed a number of CCP donors with no detectible neutralizing activity or who lost activity over time, presenting a potential problem for immunological protection and herd immunity. More extensive longitudinal studies are needed to clarify the role of T-cell based immunologic memory following infection and its relationship to antibody-based immunity. Nelde et al. observed T cell responses in over half of IgG negative convalescent individuals, and other reports are mixed on whether T cell responses correlate with circulating antibody levels [42, 43]. In the current study, we present a longitudinal serological assessment of 202 CCP donors up to 9 months post-recovery from generally mild infection. Anti-SARS-CoV-2 antibodies were identified in 97% of CCP donors at initial presentation. IgG and neutralizing antibodies were positively correlated with age, BMI and clinical severity, and persisted for up to 9 months post-recovery from infection. IgG anti-SARS-CoV-2 levels decreased below initial level in 56% of repeat donors; however, seroreversion was uncommon. Donors with low initial neutralizing antibody titers < 1:80 were more likely to lose neutralizing activity over time compared to those with initial titers > 1:80, suggesting neutralizing activity early in convalescence may be predictive of long-term persistence. Our data suggest that immunological memory is acquired in most individuals infected with SARS-CoV-2 and is sustained in a majority of patients for up to 9 months after recovery.

## Data Availability

N/A

## Acknowledgements

The authors thank NIH blood donors for their participation in the study. This work was supported by the Intramural Research Program of the NIH Clinical Center (project Z99 CL999999).

This project has been funded in whole or in part with Federal funds from the National Institute of Allergy and Infectious Diseases (NIAID), National Institutes of Health (NIH), U.S. Department of Health and Human Services (DHHS), under Contract No. HHSN272201800013C. R.S.B.,

## Author Contributions

K.W. conceived, designed and supervised clinical protocols. V.D.G conceived, designed and supervised the research study. ANH performed statistical analysis and generated all paper figures. L.C., S.P. and T.S. recruited participants, executed clinical protocols and collected clinical data. JT provided oversight to clinical sample processing and testing. R.G., J.L. and ENP performed the neutralization assays. M.R.H. provided oversight to neutralization work experiments and critically reviewed the manuscript. V.D.G., K.W., A.N.H and L.C. wrote and critically reviewed the manuscript with input from all co-authors. HJA and CCC critically reviewed the manuscript.

## Conflicts of Interest

The authors have no relevant conflicts of interest to disclose.

**Extended Data Fig. 1 – Correlation of serology tests with symptom resolution and clinical characteristics. a**. Correlation of total antibody levels (top) and IgG antibody levels (bottom) with days post-symptom resolution (n=199; excludes n=3 asymptomatic donors). Solid line represents a linear fit of the data including 95% CI (dotted lines). Pearson correlation coefficient (r) is displayed along with p-value. **b**. Distribution of days post-symptom resolution based on FRNA_50_ neutralizing antibody titers (n=199; excludes n=3 asymptomatic donors). Boxplots indicate median value with 1^st^ and 3^rd^ quartiles, and bars span minimum and maximum values. Individual patient values are indicated by dots. A statistically significant difference between groups was determined via Kruskal-Wallis test (p-value = 0.0033). Post hoc comparisons were calculated using Dunn’s multiple comparison test, with significant pairwise comparisons indicated on the graph. Total antibody levels, IgG antibody levels, and neutralizing antibody titers in CCP donors are shown based on **c**. sex (Female, n=110; Male, n=92), **d**. HLA status in females (Negative, n=81; Positive, n=26; status not determined for n=3 donors), **e**. blood type (A-, n=12; A+, n=60; AB-, n=1; AB+, n=14; B-, n=1; B+, n=25; O-, n=11; O+, n=68; unknown (UNK), n=10), and **f**. race (Asian, n=15; Black, n=14; Caucasian, n=153; Hispanic, n=9; Mixed Race, n=6). For total antibody and IgG antibody graphs, boxplots display the median with 1^st^ and 3^rd^ quartiles, and bars indicate the 5^th^ and 95^th^ percentile. Dots indicate outliers. For neutralizing titer graphs, boxplots display the median with 1^st^ and 3^rd^ quartiles, and bars span minimum and maximum values. Individual donor values are indicated by dots. In c and d, statistics calculated via Mann-Whitney test. In e and f, statistics calculated via Kruskal-Wallis test, and no significant difference between groups was observed. S/Co, signal to cutoff ratio; ns, not significant.

**Extended Data Fig. 2 – SARS-CoV-2 neutralizing titers in CCP donors over time. a**. IgG antibody levels are plotted relative to days post-symptom resolution for repeat CCP donors (n=103; n=2 asymptomatic donor not plotted). Samples from the same donor are connected by lines. Colors indicate antibody level outcomes over time: increase (green), decrease (red), no change (percent change <1%; purple), and negative (levels consistently below or falling below cutoff threshold S/Co <1; black). The S/Co threshold of 1 is indicated by the dashed line, and individual values falling below this threshold are represented by an “x”. **b**. Neutralizing titers over time for repeat CCP donors are stratified and plotted individually based on the titer recorded at first donation. Outcomes over time are quantified in the pie charts, and individual donor titers over time are plotted in the line graphs. In the line graphs, neutralizing titers are plotted relative to the number of days since the 1^st^ donation. Samples from the same donor are connected by lines. Colors indicate neutralizing titer outcomes over time: increase (green), decrease (red), no change/level off (titer levels either stayed the same for all timepoints or leveled off to a titer >0 for the last 2 or more donations; purple), and negative (consistently undetectable neutralizing activity or titers falling to an undetectable level over time; black).

