## Supplemental Figures for "Anti-SARS-CoV-2 Serology persistence over time in COVID-19 Convalescent Plasma Donors"

### Slide 1
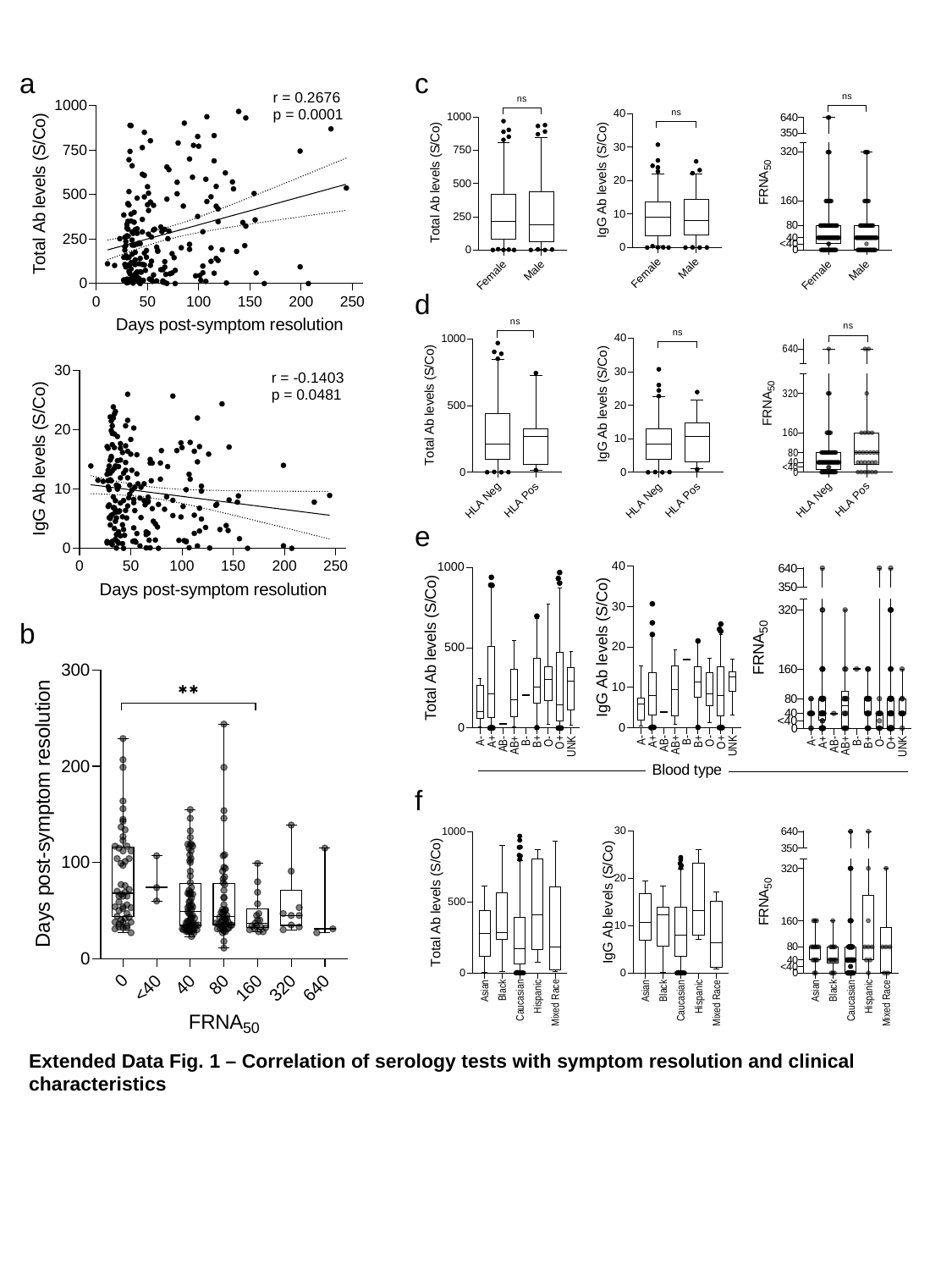

a
c
d
e
b
f
Extended Data Fig. 1 – Correlation of serology tests with symptom resolution and clinical characteristics

### Slide 2
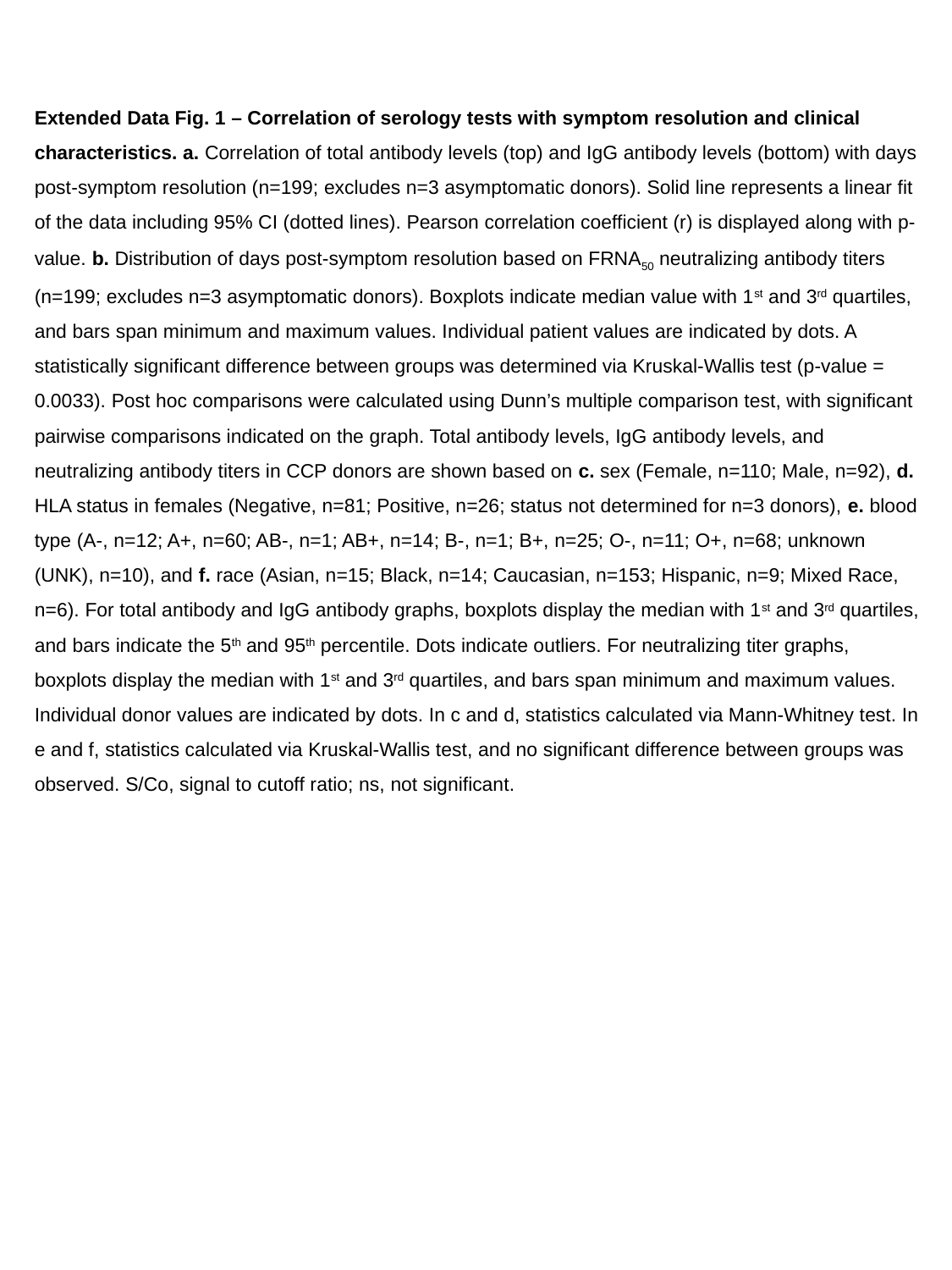

Extended Data Fig. 1 – Correlation of serology tests with symptom resolution and clinical characteristics. a. Correlation of total antibody levels (top) and IgG antibody levels (bottom) with days post-symptom resolution (n=199; excludes n=3 asymptomatic donors). Solid line represents a linear fit of the data including 95% CI (dotted lines). Pearson correlation coefficient (r) is displayed along with p-value. b. Distribution of days post-symptom resolution based on FRNA_50_ neutralizing antibody titers (n=199; excludes n=3 asymptomatic donors). Boxplots indicate median value with 1^st^ and 3^rd^ quartiles, and bars span minimum and maximum values. Individual patient values are indicated by dots. A statistically significant difference between groups was determined via Kruskal-Wallis test (p-value = 0.0033). Post hoc comparisons were calculated using Dunn’s multiple comparison test, with significant pairwise comparisons indicated on the graph. Total antibody levels, IgG antibody levels, and neutralizing antibody titers in CCP donors are shown based on c. sex (Female, n=110; Male, n=92), d. HLA status in females (Negative, n=81; Positive, n=26; status not determined for n=3 donors), e. blood type (A-, n=12; A+, n=60; AB-, n=1; AB+, n=14; B-, n=1; B+, n=25; O-, n=11; O+, n=68; unknown (UNK), n=10), and f. race (Asian, n=15; Black, n=14; Caucasian, n=153; Hispanic, n=9; Mixed Race, n=6). For total antibody and IgG antibody graphs, boxplots display the median with 1^st^ and 3^rd^ quartiles, and bars indicate the 5^th^ and 95^th^ percentile. Dots indicate outliers. For neutralizing titer graphs, boxplots display the median with 1^st^ and 3^rd^ quartiles, and bars span minimum and maximum values. Individual donor values are indicated by dots. In c and d, statistics calculated via Mann-Whitney test. In e and f, statistics calculated via Kruskal-Wallis test, and no significant difference between groups was observed. S/Co, signal to cutoff ratio; ns, not significant.

### Slide 3
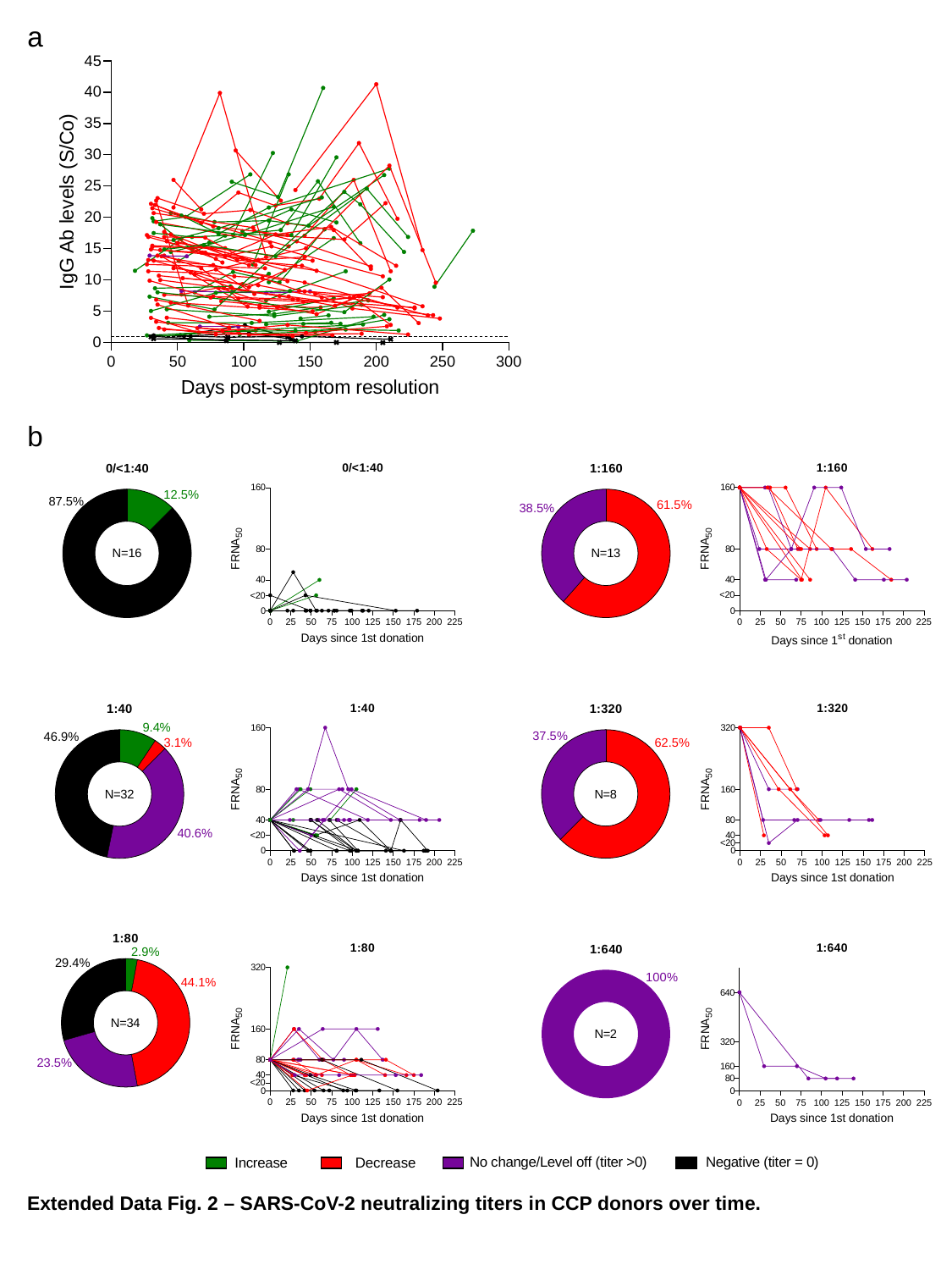

a
b
Extended Data Fig. 2 – SARS-CoV-2 neutralizing titers in CCP donors over time.

### Slide 4
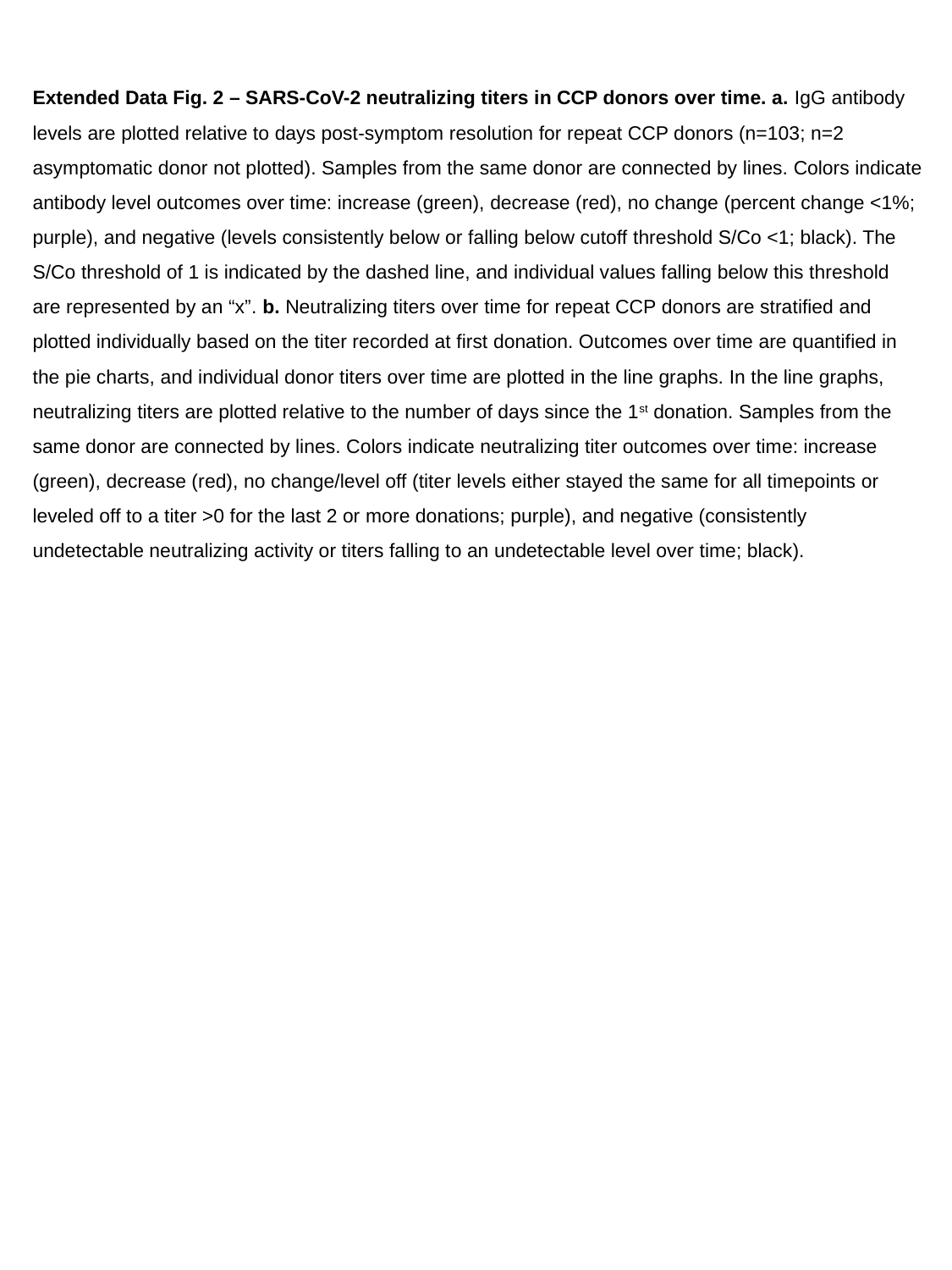

Extended Data Fig. 2 – SARS-CoV-2 neutralizing titers in CCP donors over time. a. IgG antibody levels are plotted relative to days post-symptom resolution for repeat CCP donors (n=103; n=2 asymptomatic donor not plotted). Samples from the same donor are connected by lines. Colors indicate antibody level outcomes over time: increase (green), decrease (red), no change (percent change <1%; purple), and negative (levels consistently below or falling below cutoff threshold S/Co <1; black). The S/Co threshold of 1 is indicated by the dashed line, and individual values falling below this threshold are represented by an “x”. b. Neutralizing titers over time for repeat CCP donors are stratified and plotted individually based on the titer recorded at first donation. Outcomes over time are quantified in the pie charts, and individual donor titers over time are plotted in the line graphs. In the line graphs, neutralizing titers are plotted relative to the number of days since the 1^st^ donation. Samples from the same donor are connected by lines. Colors indicate neutralizing titer outcomes over time: increase (green), decrease (red), no change/level off (titer levels either stayed the same for all timepoints or leveled off to a titer >0 for the last 2 or more donations; purple), and negative (consistently undetectable neutralizing activity or titers falling to an undetectable level over time; black).
